# The effect of galvanic vestibular stimulation on postural balance in Parkinson’s Disease: A systematic review and meta-analysis

**DOI:** 10.1101/2022.03.25.22272947

**Authors:** Mohammad Mahmud, Zaeem Hadi, Mabel Prendergast, Matteo Ciocca, Abdel Rahman Saad, Yuscah Pondeca, Yen Tai, Gregory Scott, Barry M Seemungal

## Abstract

People with Parkinson’s disease (PD) experience postural imbalance, leading to considerably increased risk of falls. Galvanic Vestibular Stimulation (GVS) is postulated to modulate postural balance in humans and improve it in PD. This systematic review and meta-analysis investigate the effects of GVS on postural balance in PD.

Six separate databases and research registers were searched for cross-over design trials that evaluated the effects of GVS on postural balance in PD. We used standardized mean difference (Hedges’ g) as a measure of effect size in all studies.

We screened 223 studies, evaluated 14, of which five qualified for the meta-analysis. Among n = 40 patients in five studies (range n= 5 to 13), using a fixed effects model we found an effect size estimate of g = 0.43 (p < 0.001, 95% CI [0.29,0.57]). However, the test for residual heterogeneity was significant (p < 0.001), thus we used a random effects model and found a pooled effect size estimate of 0.62 (p > 0.05, 95% CI [– 0.17, 1.41], I2 = 96.21%). Egger’s test was not significant and thus trim and funnel plot indicated no bias. To reduce heterogeneity, we performed sensitivity analysis and by removing one outlier study (n = 7 patients), we found an effect size estimate of 0.16 (p < 0.05, 95% CI [0.01, 0.31], I2 = 0%).

Our meta-analysis found GVS has a favourable effect on postural balance in PD patients, but due to limited literature and inconsistent methodologies, this favourable effect must be interpreted with caution.

## Background

Parkinson’s disease (PD) is the second commonest neurodegenerative disease, and its prevalence is predicted to double in the next two decades (Simon, Tanner & Brundin, 2020). Falls are a significant predictor in determining the quality of life of PD patients and are one of the main causes of hospitalisation in these patients (Martignoni et al., 2004). Despite this risk, the mainstay treatment of PD (dopaminergic medications) have little effect on fall outcomes (Dhall 2013).

Almost 80% of the falls in PD can be attributed to postural instability and gait impairments (Michałowska et al., 2005). The combination of postural instability and gait disorder (PIGD) is often considered to be a single continuum of PD (Alves et al., 2006; Kotagal et al., 2016) combining postural instability with falls and freezing of gait (FOG) (Factor et al., 2011). However, postural instability with falls and FOG are increasingly considered pathophysiologically distinct disorders, both being independent predictors of falls (Pelicioni et al., 2019), suggesting that posture and gait are mediated by distinct neuroanatomical pathways (Factor et al., 2011). This distinction further extends to objective assessments where gait measures are shown to improve in response to Levodopa treatment contrary to static postural balance measures, which get worse (Horak et al., 2016), suggesting that independent mechanisms mediate gait (dynamic balance) and postural balance.

Recently, there has been interest in using non-invasive brain stimulation techniques for modulating behaviour in healthy controls and patients. Transcranial direct-current stimulation (tDCS) has been used in PD to improve postural balance and dynamic balance (Beretta et al., 2020; Lee & Kim, 2021; Orrù et al., 2019; Scinicariello et al., 2001). However, studies have reported mixed findings, further confirmed by a recent meta-analysis showing inconclusive evidence for the impact of tDCS on postural and dynamic balance in PD (Liu et al., 2021). Moreover, the underlying mechanism of action of tDCS for modulating the postural balance remains unclear.

Galvanic vestibular stimulation (GVS) is another form of non-invasive brain stimulation, in which an electrical current is delivered to the vestibular afferents through the electrodes placed over the mastoids. GVS can cause a standing subject to sway in different directions depending on the polarity of electrodes (Scinicariello et al., 2001). GVS can be used in supra-threshold manner, in which the stimulation level is higher than the cutaneous perceptual threshold, i.e., subjects can perceive being stimulated through non-vestibular sensory afferents (Scinicariello et al., 2001; Wardman et al., 2003). Alternatively, GVS can also be used in sub-threshold manner, in which stimulation is imperceptible to subjects, i.e., lower than the cutaneous threshold (Schniepp et al., 2018; Wuehr et al., 2016). Sub-threshold GVS has been used in clinical studies and has been most useful to enhance postural balance during steady standing in young (Goel et al., 2015) and elderly healthy volunteers (Fujimoto et al., 2016), and in patients with bilateral vestibular failure (Wuehr, Decker & Schniepp, 2017). The specific type of GVS depends on the stimulation parameters such as types of waveform (e.g. Direct current (DC), sinusoid (AC), random noise), frequency band of the stimulation waveform (e.g. narrow band (0-30 Hz), broadband (white-noise)), or the electrode configuration (e.g. monopolar or bipolar) (Lee et al., 2021).

Several studies have tested sub-threshold GVS as a potential treatment to improve postural balance in PD. GVS can modulate the activation of the vestibular afferents (Fitzpatrick & Day, 2004; Wardman & Fitzpatrick, 2002), which connect to thalamus and basal ganglia (Cai et al., 2018; Stiles & Smith, 2015) and further have projections to the pedunculopontine nucleus (PPN) (Cai et al., 2018; Visser & Bloem, 2005). The link of PPN with postural balance has been established from linking severity of balance impairment to cell loss within the PPN (Hirsh et al.,1987) to stimulation studies in humans (Yousif et al., 2016). There is accumulating evidence showing cholinergic PPN neuronal loss, and not dopaminergic, is associated with worsening postural balance (Bohnen et al., 2012; Bohnen & Albin, 2009; Müller et al., 2013) but not gait (Müller et al., 2013). Further, the PPN is highly vestibular responsive (Aravamuthan & Angelaki, 2012; Yousif et al., 2016), suggesting that the PPN may mediate vestibular-dependent mechanisms of postural balance. A previous study showed that GVS can modulate PPN connectivity in PD (Cai et al., 2018) suggesting that GVS likely mediates postural balance through activation of vestibular afferents to PPN thalamus connections.

The potential promise of GVS as a treatment for reducing falls risk in PD through modulating postural balance is still unproven. However, recent studies show promise as a possible clinical application and a non-invasive neuro-modulatory treatment for PD. Thus, the aim of this meta-analysis was to assess whether current evidence shows any impact of GVS on postural balance in patients with PD.

## Methods

### Eligibility

#### Inclusion criteria

##### Types of Studies

We included only cross-over trials that evaluated the effects of GVS on postural balance in patients with PD. We excluded any conference proceedings or incomplete trials.

##### Types of Participants

We included trials that specified patients with PD as diagnosed by the UK Parkinson’s Disease Bank or an equivalent clinical diagnosis regardless of time since diagnosis, medication status, age, or Hoehn and Yahr (H&Y) stage. We excluded trials that did not specifically focus on PD.

##### Types of Intervention

We included trials if they used GVS in patients with PD. The ‘placebo’ group is defined as a sham GVS group tested pre and post sham. GVS was included regardless of type of stimulation characteristics (direct current, alternating current, random noise, frequency bandwidth, intensity, electrode arrangement/size and correlation.)

##### Exclusion criteria

Review articles, case reports, study protocols, trials assessing conditions other than postural balance, or interventions other than GVS. We furthermore excluded studies that did not use GVS but alternative methods of non-invasive brain stimulation such tDCS, transcranial alternate current stimulation (tACS), or transcranial magnetic stimulation (TMS).

### Outcome Measures

The primary outcomes were any form of postural balance assessments while standing. We included the following postural balance measures: 1.root mean square (RMS) sway 2. Sway path 3. sway area, 4. sway velocity 5. Mean of centre of pressure (COP) 6. postural angles. In scenarios when postural balance was assessed on either hard or soft surfaces with eyes open or closed, we only used the eyes closed soft surface condition. Postural balance assessment during eyes closed over soft surface has for long been shown to be the most sensitive of these conditions for identifying poor balance in patients (Black et al., 1988; Kantner et al., 1991; Cronin et al., 2017)

### Electronic Search

We performed our systematic review and meta-analysis based on criteria given by the PRISMA statement (See Appendix). Our search strategy searched the following databases: MEDLINE (from 1993 to 20 July 2020), Web of Science Core Collection, AMED (from 1993 to 1 Jan 2020), Cochrane Central Register of Controlled Trials, Google Scholar (from 2004 to 10 April 2020), The Physiotherapy Evidence Database and Rehabdata. We conducted a comprehensive search strategy to focus on peer-reviewed articles that reported an effect of GVS on a postural balance in PD patients. We adapted our search strategy according to the database (see Appendix). We identified and searched the following ongoing trial and research registers: current controlled trials, clinicaltrials.gov, European Union Clinical Trials Register, World Health Organization International Clinical Trials Registry Platform.

### Data collection and analysis

#### Selection of studies

Two review authors (MM and MP) read through the titles and abstracts identified in the electronic search. Studies were then selected according to the inclusion criteria, detailed above. Two review authors (MM and MP) then individually read through the full papers and selected those studies that were relevant according to our exclusion criteria.

#### Data extraction

From the selected studies, three authors (MM, ZH, MP) independently extracted the relevant data from the same studies. A standardised data extraction table was designed and included information about the following:

Study parameters: descriptive data, blinding, study-design, control condition

Participant characteristics: numbers, PD staging (H&Y, disease duration), medication dopamine status

GVS parameters: type of noise, intensity, frequency bandwidth, duration, correlation, electrode (location, size and arrangement)

Results: sample size, mean and variances pre- and post-GVS, mean difference

#### Assessment of Risk of Bias and trial quality

We used the Risk of Bias 2 (RoB) for crossover trials using the Cochrane Collaboration Tool (Sterne et al., 2019). For each study, MM, ZH rated the risk of selection bias, performance bias, detection bias, attrition bias, reporting bias, and other biases. These were rated from low, high or uncertain based on the guidance provided by Cochrane.

#### Dealing with missing information

In the case of any missing information, we contacted the authors of the relevant studies with the details of the data required. If we were unable to receive a response or to receive the missing data that was required for analysis, we excluded this study.

#### Meta-analytic techniques and statistical analyses

We used standardized mean difference (Hedges’ g) as a measure of effect size in all studies. Effect size calculations were performed using the method for within-subject study designs (Borenstein et al., 2021). A positive effect size denotes improvement in measured parameters in response to stimulation and vice versa for a negative effect size. A Pearson’s correlation coefficient (r), required for the effect size calculations of paired group (pre-post comparisons) designs, was calculated where raw data was available. If neither correlation coefficient was reported nor raw data was available, then correlation coefficient was imputed based on raw data available from the other studies included in the analysis.

The meta-analysis was performed in JASP software (Love et al., 2019). Pooled effect-size was first estimated using a fixed-effects model. The heterogeneity between each group was tested using the I^2^ test in JASP. Greater than 50% and significant residual heterogeneity (p < 0.05) was considered high heterogeneity. A random-effects model was applied if high heterogeneity was observed. Sensitivity analysis was performed if there was still considerable heterogeneity, and to control for any imputed values.

## Results and Discussion

### Study Classification

Figure 1 shows the number of studies screened, included, and excluded. Our search resulted in 275 hits and after removal of duplicates, 223 were screened. 55 full-length articles were assessed and after excluding any conference proceedings, irrelevant study designs/interventions/patients, we were left with 14 articles. Of these, five were eligible and included in the meta-analysis (Samoudi et al, 2014, Pal et al., 2009, Tran et al., 2018, Katoaka et al., 2016, Okada et al., 2015).

**Figure 1.**
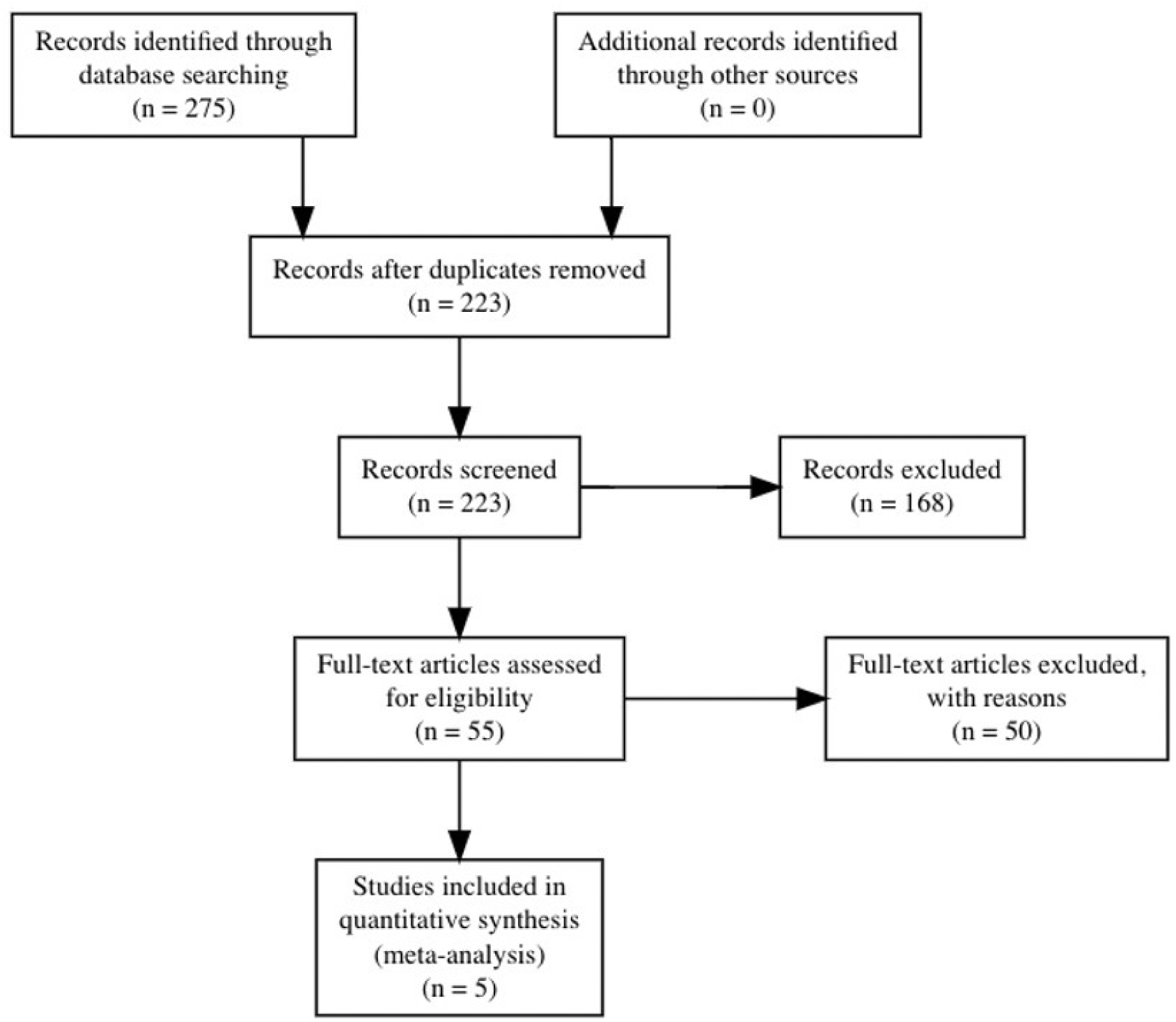
PRISMA flow diagram – From a total of 275 identified research article using the appropriate screening and inclusion criteria the synthesis included five studies in total

#### Study design and participant characteristics (Table 1 and 2)

**Table 1.** Study design, stimulation dosage and outcome measures used in the included studies.

| **Study** | **Study Design** | **Dosage** |  | **Outcomes** |
| --- | --- | --- | --- | --- |
|  |  | **Trials of verum condition** | **Time between trials** |  |
| Pal et al.<br>(2009) | Single-blinded<br>Non-randomised<br>Sham-controlled | 6 trials of each intensity and condition (24 total) | No data | Mean centre of pressure (Anterior-Posterior & Medial-lateral direction) |
| Katoaka et al. (2016) | Double-blinded<br>Randomised<br>Crossover<br>Sham-controlled | 1 trial | 1 week between verum and sham | Anterior trunk bending angle |
| Samoudi et al.<br>(2014) | Double-blinded<br>Randomised<br>Crossover<br>Sham-controlled | 1 trial | Crossover performed on separate days, time | Sway (sway-path) |
| Okada et al. (2015) | Single-blinded<br>Randomised<br>Sham-controlled | 1 trial | 1 week<br>between<br>crossovers | Anterior bending angles |
| Tran et al. (2018) | Single-blinded<br>Randomised<br>Sham-controlled | 4 trials | 60 seconds<br>rest between<br>trials | Mean centre of<br>pressure (Anterior-Posterior & Medial-lateral direction) |

**Table 2.**
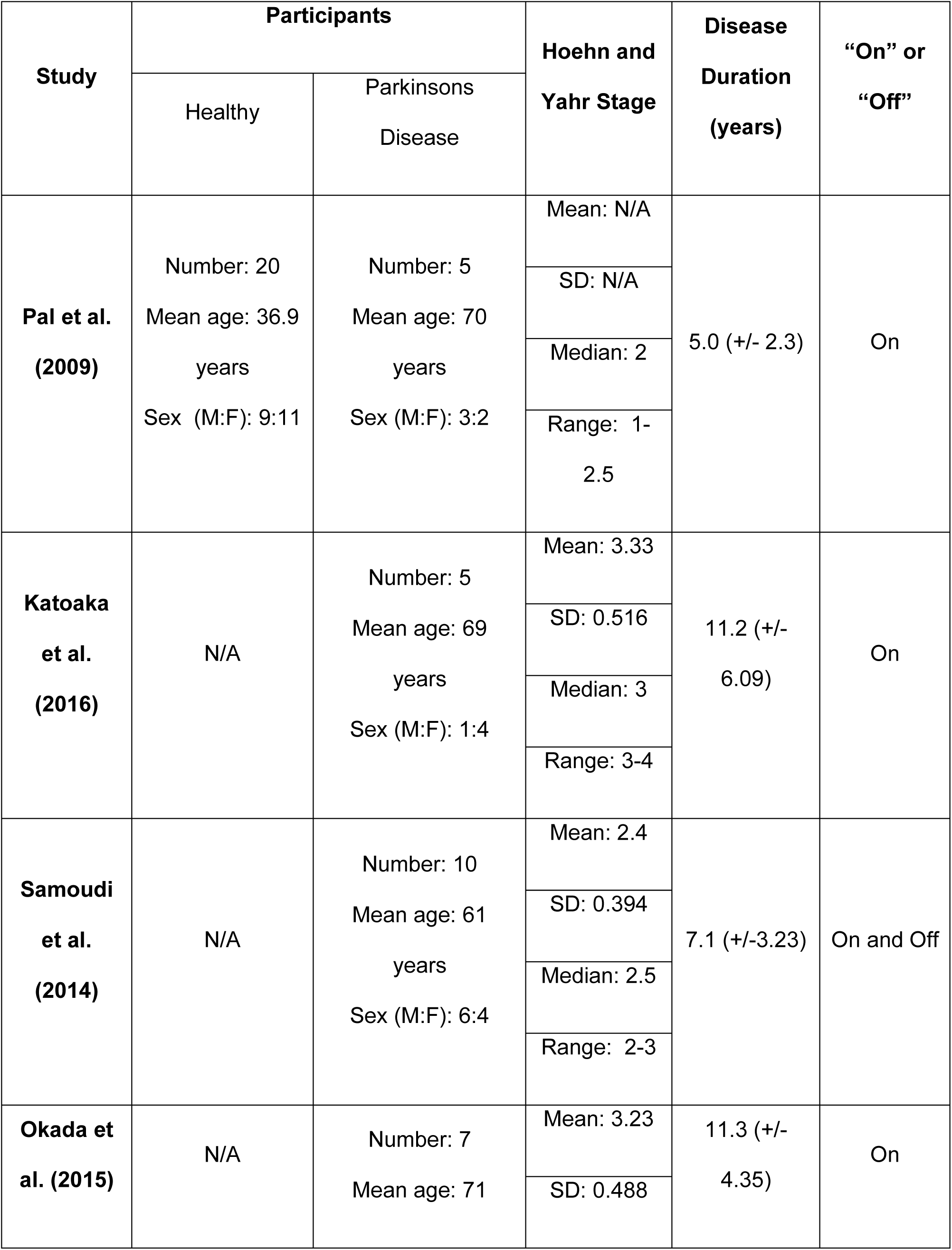
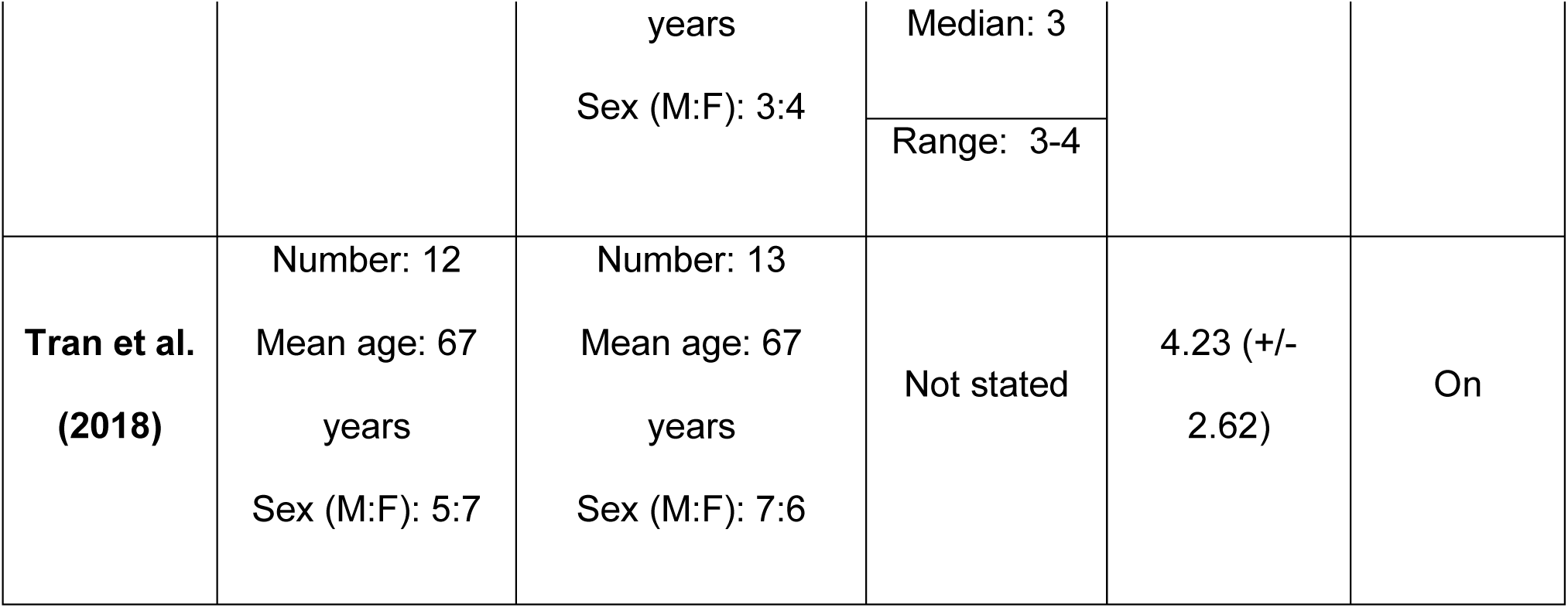
Participant Characteristics including if the study had a comparison control group and stage of Parkinson’s diasese, years since diagnosis and if study was conducted during on or off phases.

##### Stimulation characteristics

A total of n = 40 subjects were included in the analysis from five studies (Table 3) (Samoudi et al., 2014; Pal et al., 2009; Tran et al., 2018, Katoaka et al., 2016, Okada et al., 2015). One (Pal et al., 2009) of the five studies used pre-fixed stimulus intensities of 0 mA, 0.1 mA, 0.3 mA, and 0.5 mA. Two of five studies (Kataoka et al., 2016; Okada et al., 2015) had similar protocols in which GVS stimulation was ramped up slowly to 0.7 mA and then stimulation was applied for 20 minutes. The remaining two studies used stimulation intensities based on a subject’s individual cutaneous perceptual thresholds (Samoudi et al., 2015; Tran et al., 2018).

**Table 3.**
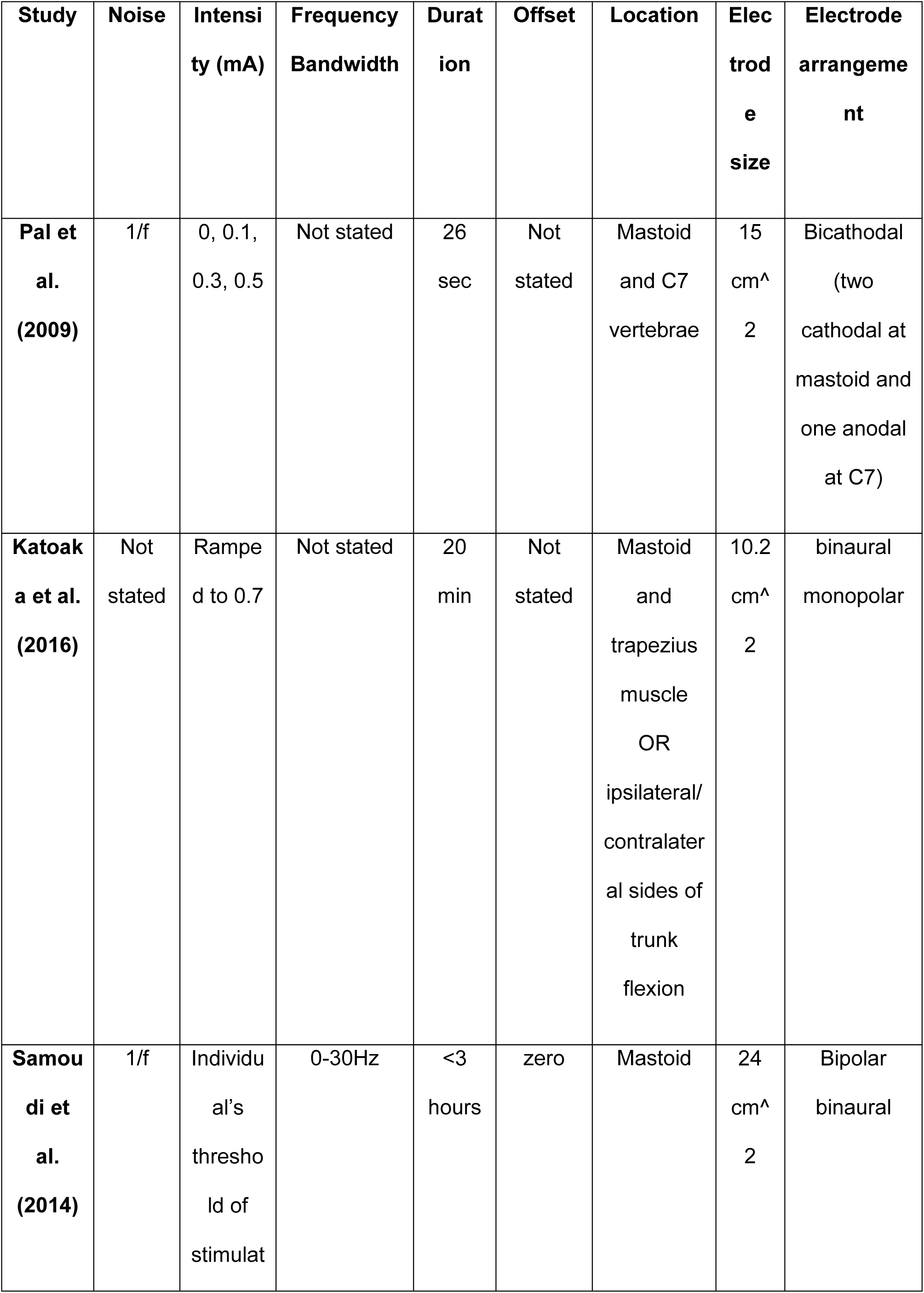
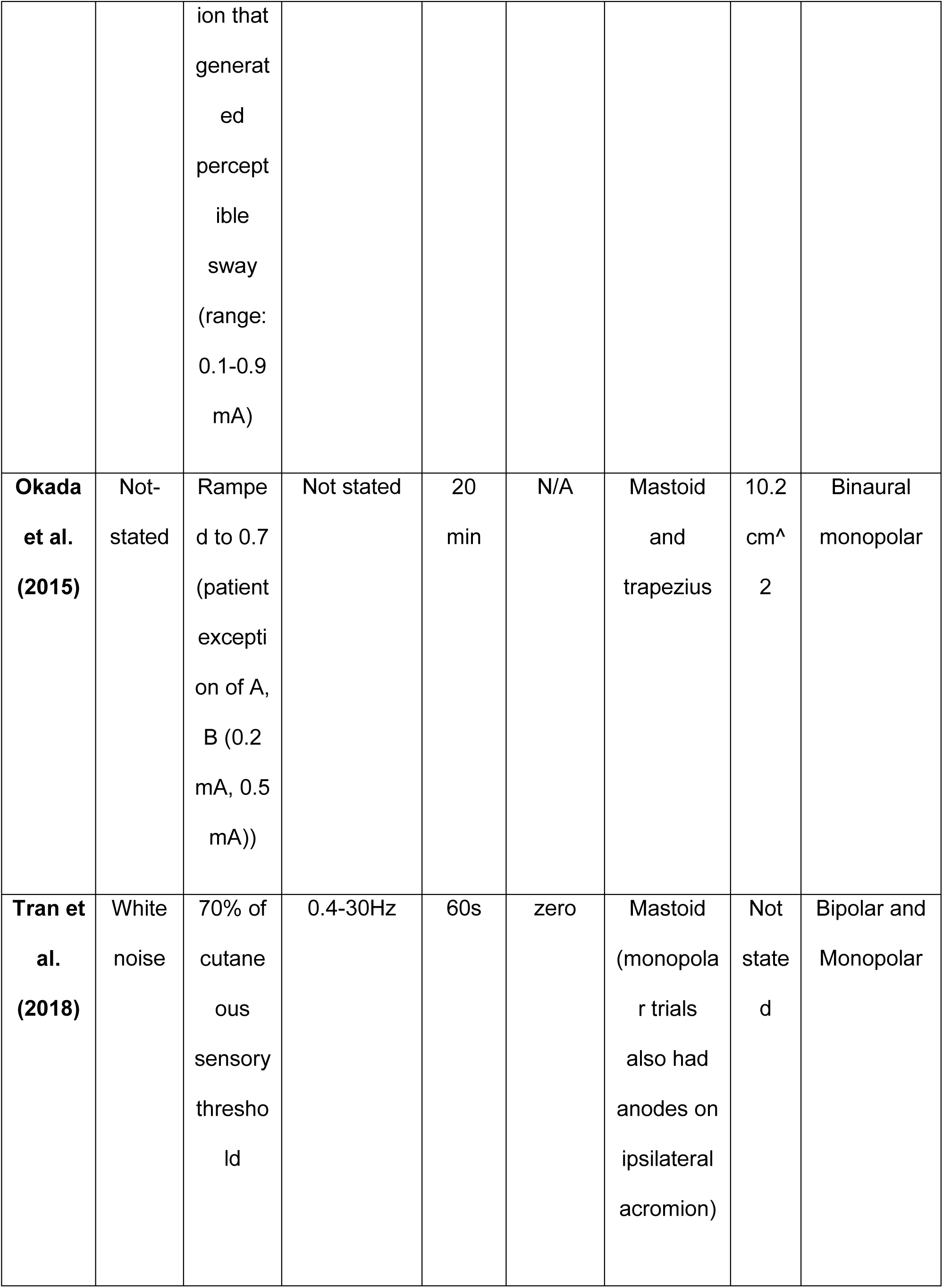
Stimulation Characteristics

All studies involve electrode placement over the mastoid processes, however, one study used two cathodes over mastoids with anode over the C7 vertebrae (Pal et al., 2009) and two studies also stimulated trapezius muscles in addition to mastoids (Kataoka et al., 2016). Two of the studies (Okada et al., 2015; Tran et al., 2018) used monopolar stimulation, two used bipolar (Kataoka et al., 2016; Samoudi et al., 2015), and one (Pal et al., 2009) used bicathodal stimulation over the mastoids. The duration of the stimulation was 26 seconds (Pal et al., 2009), 60 seconds (Tran et al., 2018), 20 minutes (Kataoka et al., 2016; Okada et al., 2015), and up to 3 hours (Samoudi et al., 2015).

One (Pal et al., 2009) of the five included studies did not specify the frequency band of the random noise in noisy GVS. Two studies (Kataoka et al., 2016; Okada et al., 2015) did not mention the choice of frequency band, possibly due to the use of Direct Current GVS, however, it was not explicitly mentioned. One study used white noise in 0.4-30 Hz band (Tran et al., 2018) and one other used 1/f noise in 0-30 Hz band (Samoudi et al., 2015).

All five studies used PD patients who were assessed during the ON medication condition, while the OFF condition was also present in some studies. However, for the sake of homogeneity, only the results for the ON condition are included.

##### Methodological Quality

RoB for crossover studies was used to assess the quality of RCTs and assess methodological areas for concern. Four of the five included studies were randomised, however no study mentioned the method of randomisation. There were no concerns identified by RoB due to carry over effects in four studies and some concern in one study (Figure 2. DS). Two studies had some concern due to deviation from intended intervention while three studies were low risk (Figure 2. D2). None of the studies had missing outcome data and thus all were considered low risk (Figure 2. D3). Four studies had low concern for outcome measurement whereas one study had some concern (Figure 2. D4). Two of the studies were considered high risk whereas three considered low risk for reporting correct results and pre-specifying the analysis performed (Figure 2. D5).

**Figure 2.**
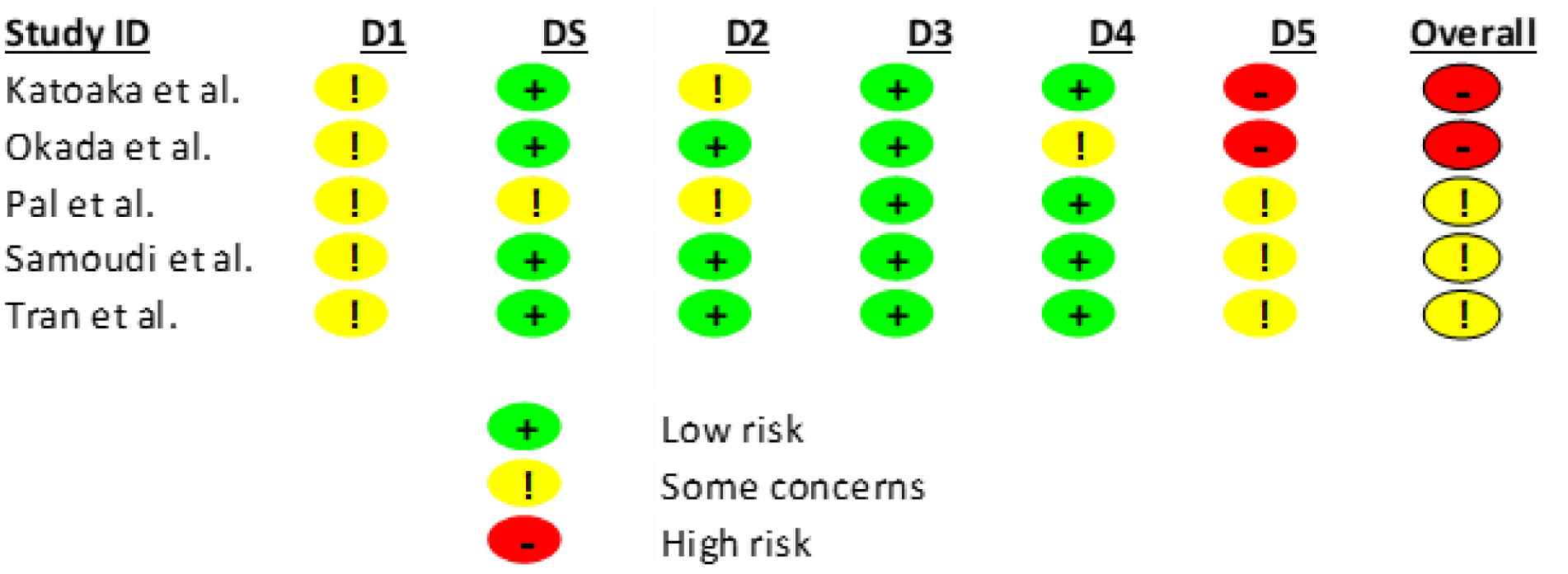
The Cochrane’s risk of bias summary; review authors’ judgements about each risk of bias item for each included study. This suggests overall concerns or high risk for all studies. D1 – Randomisation process, DS Bias arising from period and carryover effects, D2 Deviations from intended interventions, D3 Missing outcome data, D4 Measurement of the outcome, D5 Selection of the reported result.

##### Overall effect of GVS on postural balance

Using a fixed-effects model, we found an effect size estimate of g = 0.43 (p < 0.001, 95% CI [0.29,0.57]) (Figure 3), with test for residual heterogeneity indicating significant heterogeneity between studies (p < 0.001). We thus used a random effects model and found the pooled effect size estimate to be 0.62 (p > 0.05, 95% CI [– 0.17, 1.41], I^2^ = 96.21%) (Figure 4). The Egger’s test was not significant and thus a funnel plot (Figure 5) indicated no bias, however, heterogeneity was considerably high (I^2^ = 96.21%). The high heterogeneity was not unexpected considering the presence of a study with a large effect size (Hedge’s g = 2.19) and small sample (Okada et al., 2015) in the pooled effect size estimate. To reduce heterogeneity, we performed sensitivity analysis and, by removing the outlier study (Okada et al., 2015) with n = 7, we found the effect size estimate to be 0.16 (p < 0.05, 95% CI [0.01, 0.31], I^2^ = 0 %), shown in Figure 6. Note that the Egger’s test was still not significant, suggesting the funnel plot indicated no publication bias after removal of the outlier (Figure 7).

**Figure 3.**
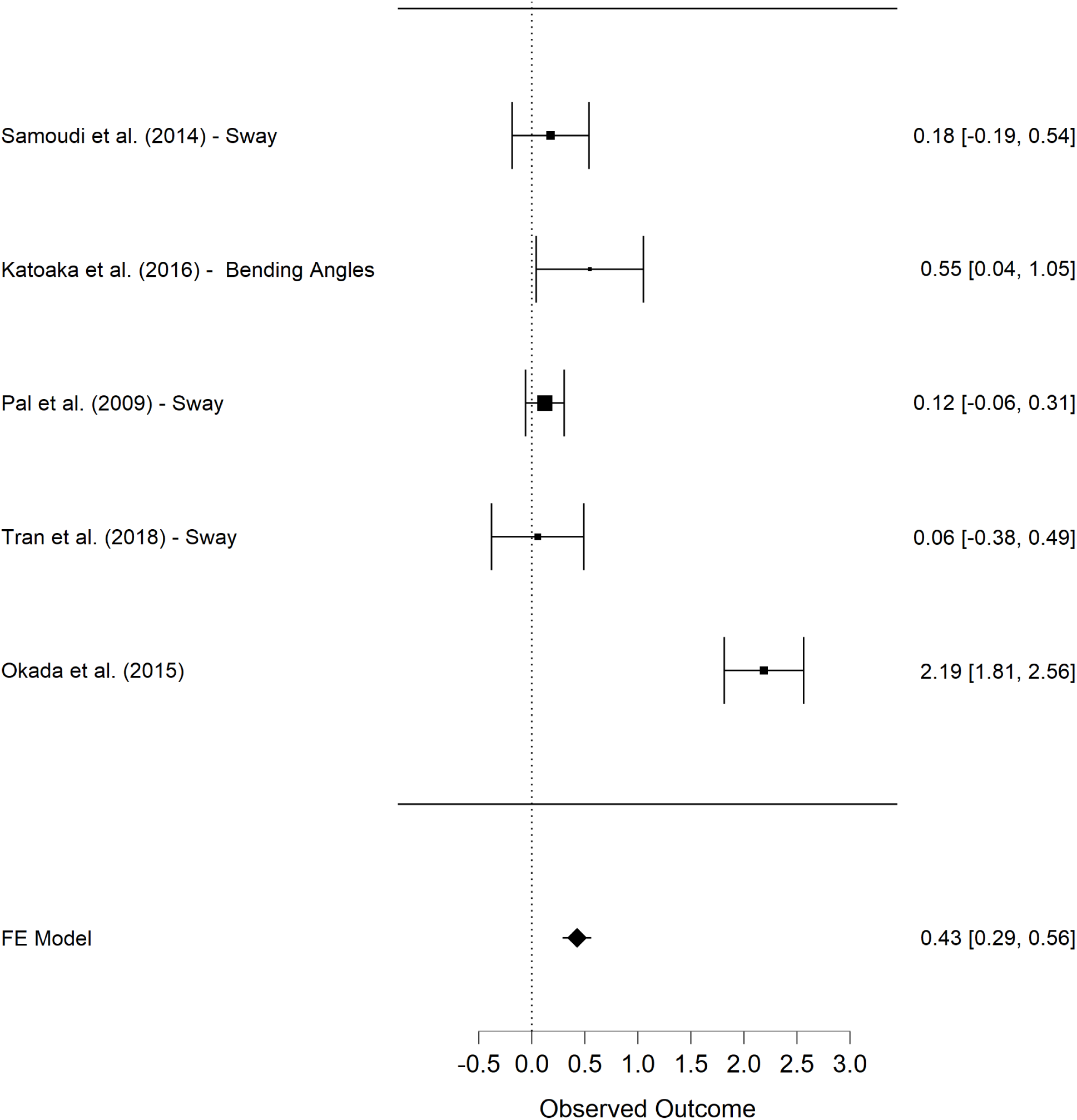
Forest Plot for Fixed Effects Model. Pooled effect size estimate using fixed effects model was g = 0.43 (p < 0.001, 95% CI [0.29,0.57]).

**Figure 4.**
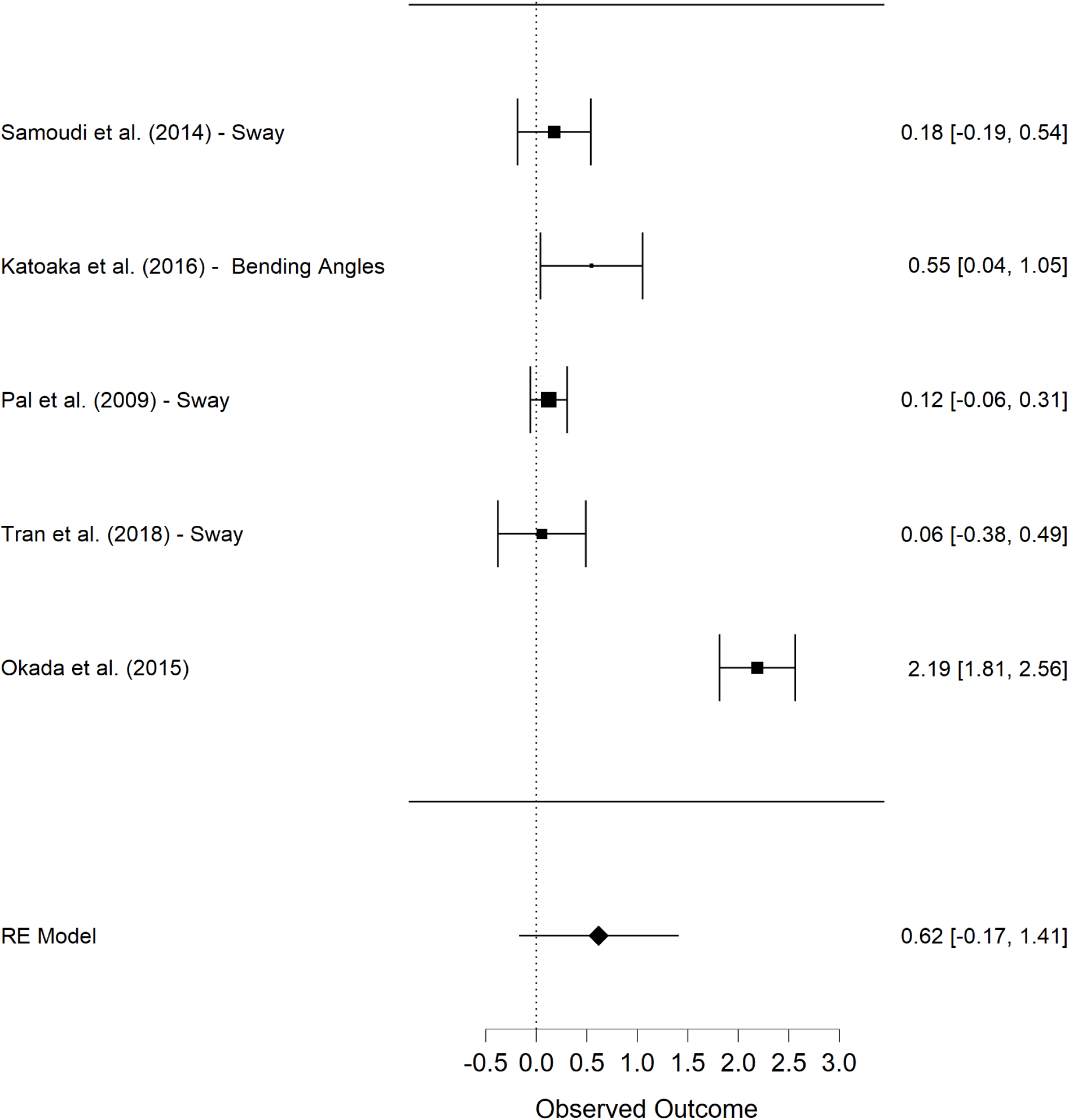
Forest Plot for Random effects model. Pooled effect size estimate using random effects model was 0.62 (p > 0.05, 95% CI [– 0.17, 1.41], I^2^ = 96.21%).

**Figure 5.**
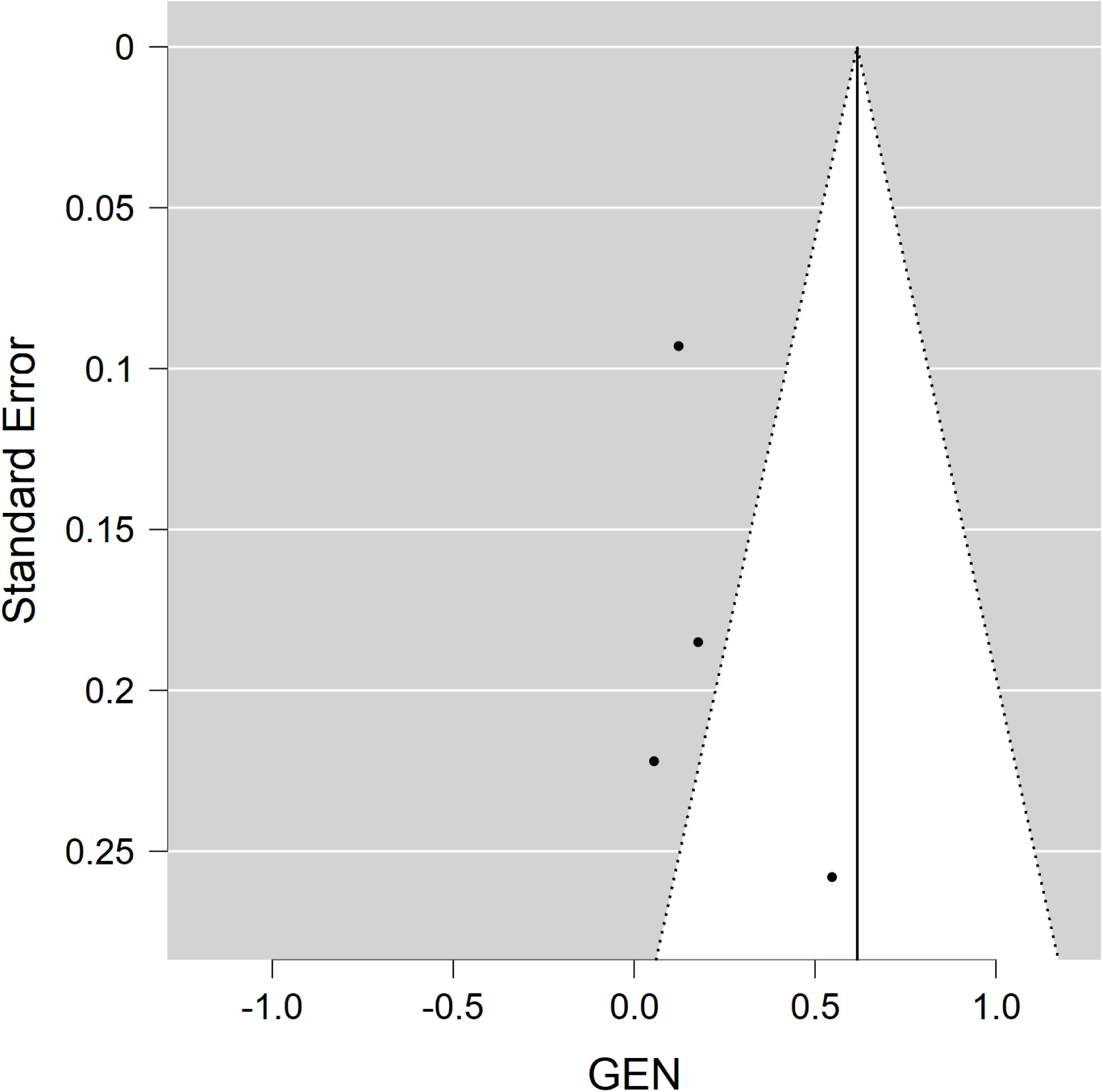
Funnel plot. The Egger’s test was not significant (p < 0.05), which indicates no bias identified via funnel plot. However, one of the studies (Okada et al., 2015) had quite large effect size (Hedge’s g = 2.19), thus adding a visually observable publication bias in the funnel plot.

**Figure 6.**
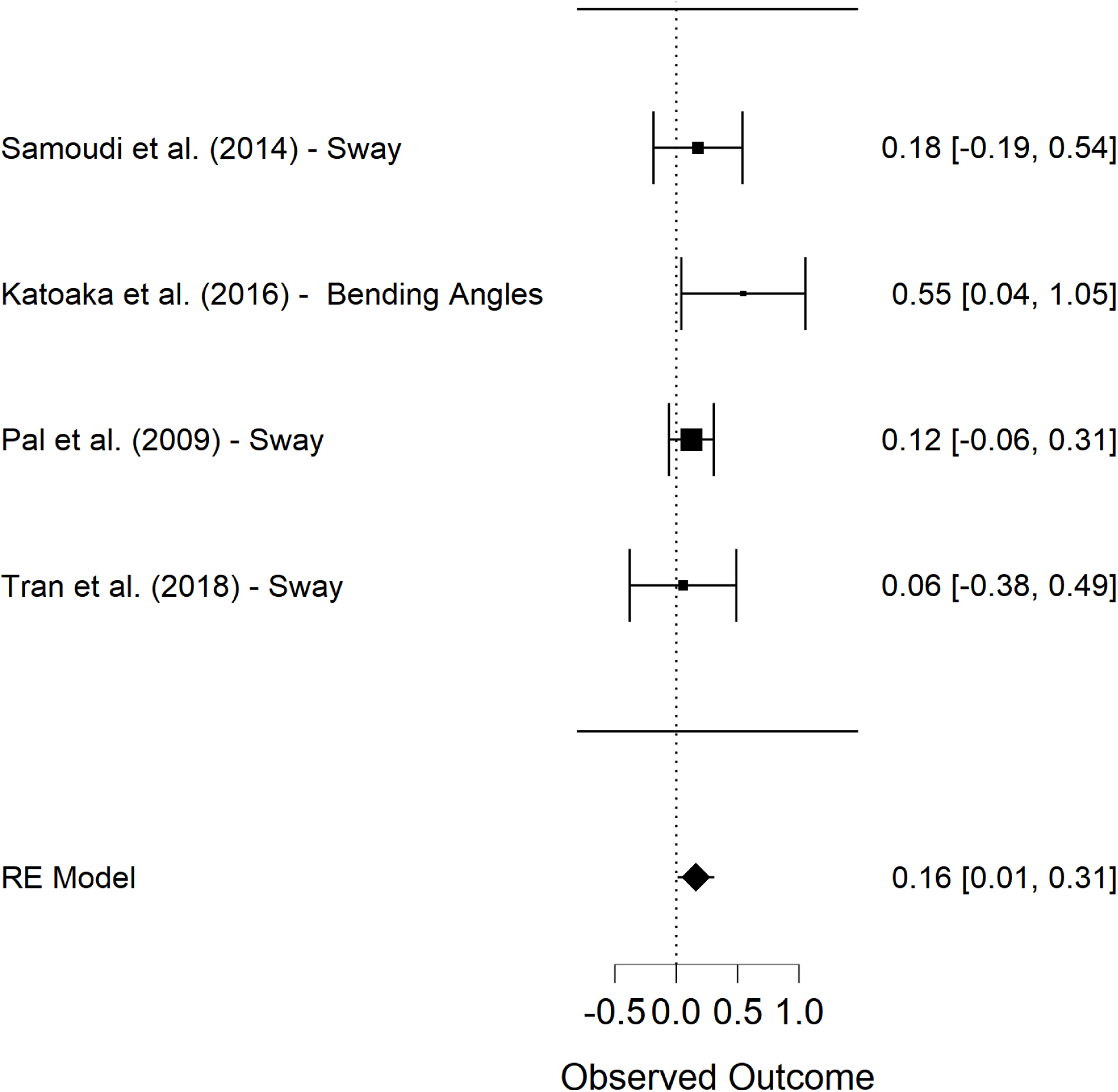
Forest Plot after Outlier Removal. After removal of the outlier study (Okada et al., 2015), pooled effect size estimate using random effects model was 0.16 (p < 0.05, 95% CI [0.01, 0.31], I^2^ = 0 %).

**Figure 7.**
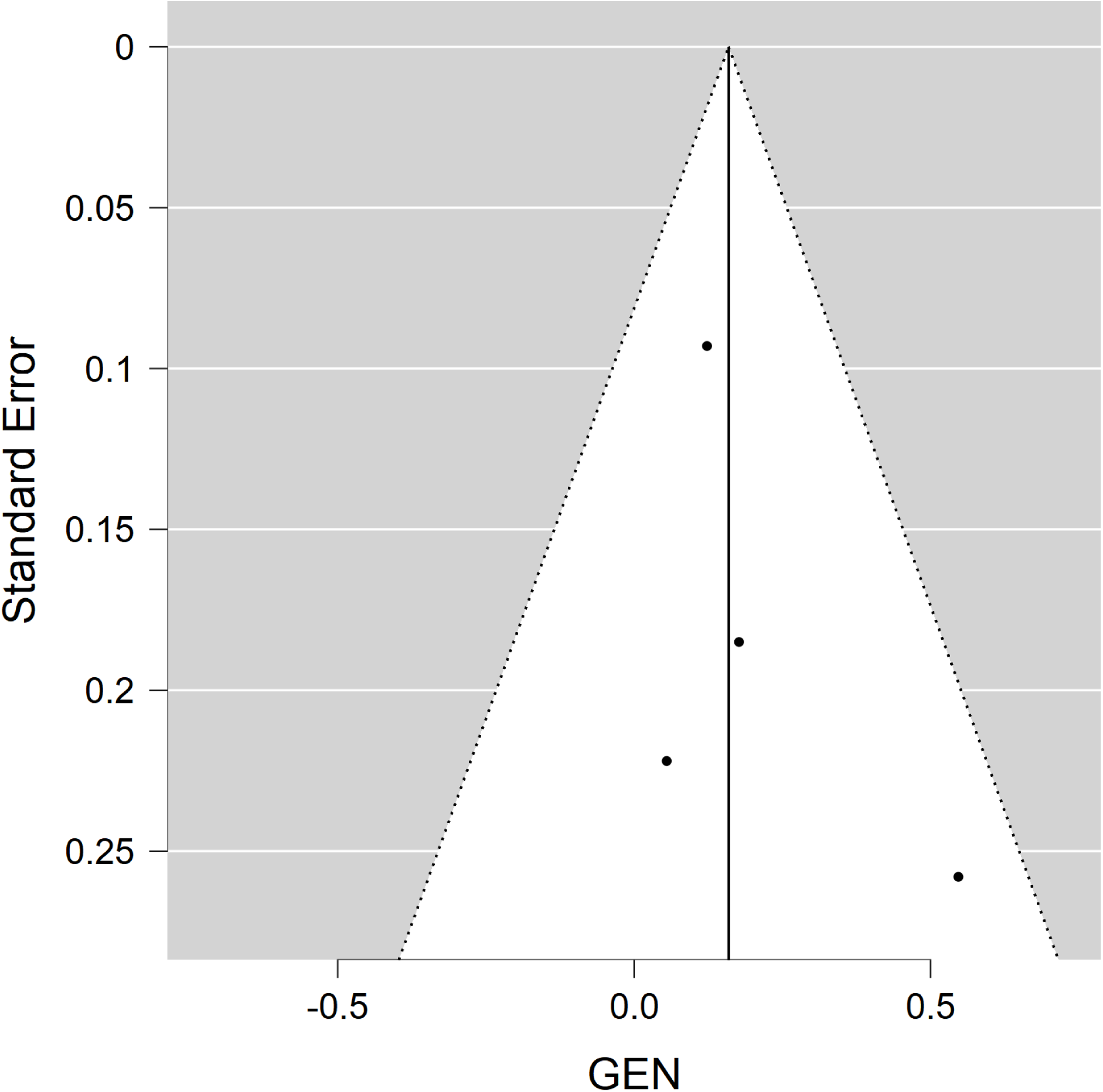
Funnel Plot after Outlier Removal. Egger’s test was not statistically significant and visual inspection of funnel plot also indicated no publication bias.

After removing the outlier study to reduce heterogeneity, there was one study (Samoudi et al., 2015) for which the correlation coefficient (*r*) was imputed (*r = 0*.*8*). However, sensitivity analysis by imputing different ‘*r’* values (*r = 0*.*6 & r = 0*.*7*) showed no change in pooled effect size estimate or its statistical significance. One study (Pal et al., 2009) used fixed stimulus intensities of 0 mA, 0.1 mA, 0.3 mA, and 0.5 mA. We selected 0.5 mA intensity for pooling effect sizes as it was closest to the other studies optimal intensities (Samoudi et al, 2014, Tran et al., 2018, Katoaka et al., 2016, Okada et al., 2015). However, to minimise bias, we performed a sensitivity analysis and found that there was little change in effect size estimate for both 0.1 mA (SMD = 0.12, p > 0.05, 95% CI [-0.08, 0.32], I^2^ = 20.95 %) and 0.3 mA (SMD = 0.15, p > 0.05, 95% CI [-0.05, 0.36], I^2^ = 0 %) and that both were not statistically significant (Figure 8 and Figure 9 respectively).

**Figure 8.**
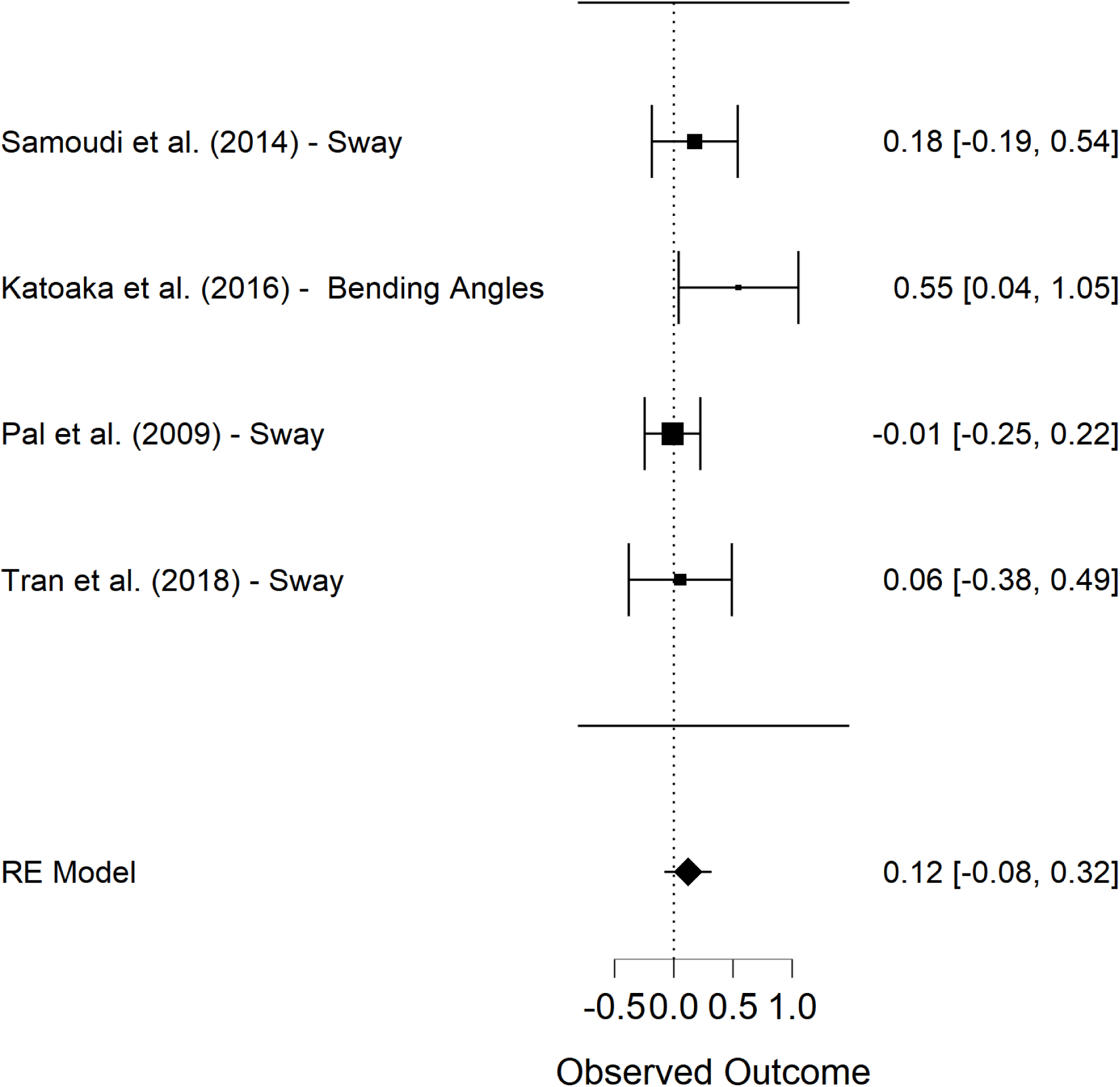
Forest Plot for Sensitivity Analysis for Pal et al., 2009. For intensity of 0.1 mA, pooled effect size estimate using random effects model was 0.12 (p > 0.05, 95% CI [-0.08, 0.32], I^2^ = 20.95 %)

**Figure 9.**
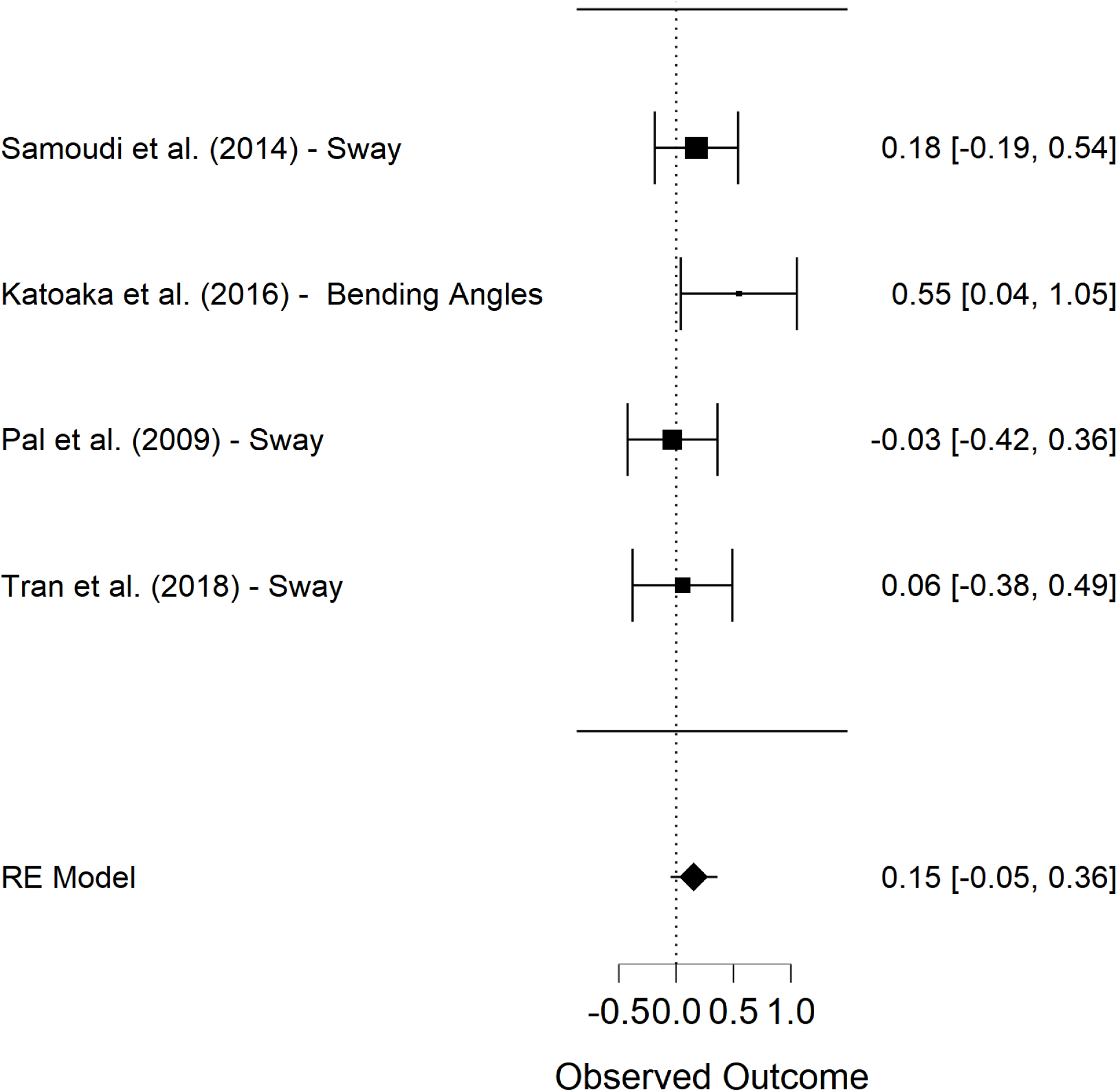
Forest Plot for Sensitivity Analysis for Pal et al., 2009. For intensity of 0.3 mA, pooled effect size estimate using random effects model was 0.15 (p > 0.05, 95% CI [-0.05, 0.36], I2 = 0 %)

## Discussion

Our meta-analysis investigated the effects of GVS on postural balance outcomes in PD patients. We found that GVS has a favourable effect on the postural balance of PD patients, however, we advise caution in interpreting our findings as this meta-analysis is limited by a small number of studies, with small sample sizes, and high heterogeneity of stimulation protocols. None of the five studies included in the meta-analysis met the correct standards of a randomised controlled trial and only two of five studies were double-blinded.

GVS protocols used between studies are inconsistent, thus decreasing the confidence in the evidence and increased heterogeneity. One (Pal et al., 2009) of the five studies used pre-fixed stimulus intensities of 0 mA, 0.1 mA, 0.3 mA, and 0.5 mA. Two of five studies (Kataoka et al., 2016; Okada et al., 2015) had similar protocol in which GVS stimulation was ramped up slowly to 0.7 mA and then stimulation was applied for 20 minutes. Whereas, the remaining two of the five studies used stimulation intensities based on subject’s individual cutaneous perceptual thresholds (Samoudi et al., 2015; Tran et al., 2018). Previous studies have shown that a subject-specific noisy GVS can improve the postural and dynamic balance in patients (Iwasaki et al., 2014; Schniepp et al., 2018) as well as healthy volunteers (Fujimoto et al., 2016; Goel et al., 2015; Mulavara et al., 2011; Wuehr et al., 2016). Similar findings have also been reported in studies using fixed stimulation intensities for all subjects (Inukai et al., 2018; Scinicariello et al., 2001; Wardman et al., 2003). It is difficult to suggest whether fixed intensity GVS or subject-specific noisy GVS is better either in healthy or any patient groups considering that both types of GVS have reported improvement in postural balance measures. From the studies included in this meta-analysis, (Samoudi et al., 2015) and (Tran et al., 2018) used nGVS with subject specific intensities but only (Samoudi et al., 2015) found improved postural balance in PD. Moreover, (Pal et al., 2009) using fixed intensity stimuli also found improved postural balance. Thus, more studies that compare stimulation types are required for better understanding.

During our sensitivity analysis, we found no impact of imputing different correlation values on the effect size estimate or its significance suggesting there was unlikely to be a bias because of imputed correlation value. One study (Pal et al., 2009) had multiple stimulation intensities, however, we selected 0.5 mA intensity for pooling effect sizes as it was closest to the other studies included in the meta-analysis (Samoudi et al, 2014, Tran et al., 2018, Katoaka et al., 2016, Okada et al., 2015). A sensitivity analysis was performed to minimise bias and we found that only 0.5 mA had resulted in overall statistically significant effect size estimate. The stimulus intensities of 0.1 mA and 0.3 mA resulted in an overall statistically insignificant effect size estimate; however, it is important to notice that actual effect sizes do not vary much with 0.1 mA (Hedge’s g = 0.12), 0.3 mA (g = 0.15), and 0.5 mA (g = 0.16).

All studies involve electrode placement over mastoid processes, however, one study used bicathodal stimulation over mastoids with anode over C7 vertebrae (Pal et al., 2009) and two studies also stimulated trapezius muscles in addition to mastoids (Kataoka et al., 2016). It’s unclear that difference in electrode positioning could have caused any impact on outcomes considering all included studies stimulated mastoids. However, contrary to the traditional stimulation configuration of bipolar electrode placement used in noisy GVS studies, two studies (Okada et al., 2015; Tran et al., 2018) used monopolar stimulation and one (Pal et al., 2009) used bicathodal stimulation over mastoids.

The duration of the stimulus also showed great variation. Some studies used durations as short as 26 seconds (Pal et al., 2009) or 60 seconds (Tran et al., 2018) whilst others used longer durations of 20 minutes (Kataoka et al., 2016; Okada et al., 2015) extending up to 3 hours (Samoudi et al., 2015). There is dispute in the literature over the long-term effects of short-duration galvanic vestibular stimulation. Some studies have noted a significant long-term effect (Kerkhoff et al., 2011, Wilkinson et al., 2014, Son, Blouin & Inglis, 2008, Hassan et al., 2021) while some studies have questioned the validity of these results due to unreliable protocols (Nooristani et al., 2019., Keywan et al., 2020).

Three (Kataoka et al., 2016; Okada et al., 2015; Pal et al., 2009) of the five included studies did not specify the frequency band of the noisy GVS and only two studies reported the type of noise as well as frequency band with one using white noise in 0.4-30 Hz band (Tran et al., 2018) and other using 1/f noise in 0-30 Hz band (Samoudi et al., 2015). The consensus on this is likely reflected by a physiological basis that low-frequency band (0-30Hz) reflect the frequency bandwidth of the vestibular system (Dlugaiczyk et al., 2019).

From the five studies included in the meta-analysis, pooled effect size was estimated from n=40 samples from five studies, which was further reduced to n = 33 when an outlier study (Okada et al., 2015) was removed to reduce the heterogeneity. From the studies included in meta-analysis, the maximum sample size of PD patients was 13 (Tran et al., 2018) with all other samples either 10 (Samoudi et al., 2015) or lower. In addition, none of the studies provided any justification of their selection of sample size or power calculations. This highlights the need for properly designed studies that fulfil all requirements of a RCT.

Another important consideration was that only two studies used control group (Table 2). It is a common practice to not compare the effect of pre-post GVS in patient cohorts with a pre-post healthy control group. Without this healthy control group comparison, the pre-post difference in patient groups may reflect only normal variance of data (Nieuwenhuis et al., 2011). Only two studies (Pal et al., 2009; Tran et al., 2018) we assessed in this review compared controls and patient groups.

Finally, one must consider how postural balance measures in PD are interpreted. Early data initially suggested that patients with PD have increased sway (Horak et al., 1992) but it seems a majority of later studies find a decreased sway at baseline (Chastan et al., 2008; Kitamura etal.,1993; Schmit et al., 2006), and some studies suggesting this increased sway does indeed correlate with increased falls risk (Frenlach et al., 2009; Rossi et al., 2009).

## Conclusion

In conclusion, the use of GVS in patients with PD shows an overall positive impact on postural balance but this review shows the evidence is inconclusive for the following reasons: 1) low statistical power, 2) heterogenous stimulation parameters and methodologies, 3) lack of appropriate randomisation. This suggests that future research into this field would benefit from appropriately powered and randomised studies that assess appropriate outcome measures of postural control in Parkinson Disease.

## Data Availability

All data produced in the present study are available upon reasonable request to the authors

## Author contributions

MM - Conceptualization; Data curation; Formal analysis; Investigation; Methodology; Project administration; Roles/Writing - original draft; review & editing. ZH - Data curation; Formal analysis; Investigation; Methodology; Validation; Visualization;; Writing - review & editing. MP - Conceptualization; Data curation; Formal analysis; Visualization; Roles/Writing - original draft. MC - Methodology, Writing - review & editing. ARS, YP, GS - Writing - review & editing. BMS - Supervision; Writing - review & editing.

## Data availability

Data and code for the meta-analysis can be found here:

## Acknowledgments

We would like to thank all authors who kindly responded to our data requests.

## Appendix

### Search strategy used for PubMed/MEDLINE and Google Scholar

(“vestibular stimulation”[tw] OR “Galvanic Vestibular Stimulation”[tw] OR “GVS”[tw] OR stochastic[tw] OR “noisy vestibular stimulation”[tw] OR “stochastic galvanic vestibular stimulation”[tw] OR “stochastic vestibular stimulation”[tw]) AND (Parkinson[tw] OR “Parkinson’s Disease”[tw] OR Parkinson’s[tw] OR PD[tw] OR IPD[tw] OR “Lewy Body Parkinson’s Disease”[tw] OR “Primary Parkinsonism”[tw] OR “Paralysis Agitans”[tw] OR Parkinson’s) AND (“randomised controlled trials” OR “randomized controlled trials” OR “random allocation” OR “controlled clinical trials” OR control* OR double-blind* OR single-blind* OR placebo OR “placebo effect” OR “cross-over” OR “cross-over study” OR therapies, investigational OR research design OR evaluation study* OR “randomized controlled trial”[pt] OR controlled clinical trial [pt] OR random*[tw] OR controlled adj5 trial* OR clinical* trial*[tw] OR cross-over[tw] OR crossover [tw] OR “Cross over”[tw] OR placebo*[tw] OR Sham[tw] OR controls[tw] OR assign*[tw] OR alternate[tw] OR allocat*[tw] OR counterbalance*[tw] OR multiple baseline[tw] OR versus[tw] OR ((control OR treatment OR experiment* OR intervention) adj5 (group* OR subject* OR patient*))[tw] OR (quasi-random* OR quasi random* OR pseudo-random* OR pseudorandom*)[tw] OR ((multicenter OR multicentre OR therapeutic) adj5 (trial* or stud*))[tw] OR ((control OR experiment* OR conservative) adj5 (treatment OR therapy OR procedure OR manage*))[tw] OR ((singl* OR doubl* OR tripl* OR trebl*) adj5 (blind* OR mask*))[tw])

#### Types of databases

##### MEDLINE

- Search strategy above
- 137 search results found
- 20 screened
- Found: Okada, katoaka, Samoudi,

##### Web of Science Core Collection

- **“**Vestibular stimulation, Parkinson’s”
- 68 results found
- Duplicates: 21
- 22 screened
- Found: Tran, Pal

##### AMED

- “Vestibular stimulation, Parkinson’s”
- 34 search results found
- Duplicates: 20
- 5 screened

##### Cochrane Central Register of Controlled Trials

- “Vestibular stimulation, Parkinson’s”
- 24 search results
- Duplicates: 5
- Screened: 5

##### Google Scholar

- Search strategy above
- 10 results
- 4 duplicates
- Screened: 3

##### The Physiotherapy Evidence Database

- **“**Vestibular stimulation, Parkinson’s”
- 1 result found
- Duplicates: 1
- 0 screened

##### Rehabdata

- **“**Vestibular stimulation, Parkinson’s”
- 1 result found
- Duplicates: 1
- 0 screened

CINAHL - No access

Inspec - No access

